# Inhibition of the beta-1 adrenergic receptor does not potentiate mirabegron-stimulated human brown adipose tissue thermogenesis

**DOI:** 10.1101/2023.03.22.23287600

**Authors:** Lauralyne Dumont, Alexandre Caron, Gabriel Richard, Etienne Croteau, Mélanie Fortin, Frédérique Frisch, Serge Phoenix, Stéphanie Dubreuil, Brigitte Guérin, Éric E. Turcotte, André C. Carpentier, Denis P. Blondin

## Abstract

Pharmacological stimulation of human brown adipose tissue (BAT) has been hindered by either ineffective activation or undesirable off-target secondary effects. Oral administration of the maximal allowable dose of mirabegron (200 mg), a β_3_-adrenergic receptor <u>(β_3_-AR)</u> agonist, has been effective in stimulating BAT thermogenesis and whole-body energy expenditure. However, this too has been accompanied by undesirable cardiovascular effects. Combining mirabegron with a cardio-selective β_1_-AR antagonist could not only suppress these unwanted effects, but potentially increase the sensitivity of the β_3_-AR and β_2_-AR in WAT and BAT. Here we report that co-ingesting a high dose of the β_1_-AR antagonist bisoprolol with mirabegron suppresses the increase in heart rate, systolic blood pressure and myocardial oxygen consumption. However, it also blunted the mirabegron-stimulated increase in BAT lipolysis, thermogenesis and glucose uptake. Whether the attenuation in BAT blood flow induced by the large dose of bisoprolol limited BAT thermogenesis remains to be determined. clinicaltrials.gov (NCT04823442)

## INTRODUCTION

The stimulation of brown adipose tissue (BAT) thermogenesis has long been touted as an attractive therapeutic target for the prevention or treatment of obesity and its cardiometabolic complications. This stems primarily from the large body of work in preclinical models, which robustly demonstrate the energy dissipating capacity of BAT following cold or pharmacological stimulation ^1–5^. However, evidence for BAT functioning in such a capacity remains controversial ^6^ and largely associative in humans ^7, 8^. Some of the barriers in understanding the therapeutic potential of human BAT relates to the stimuli used to activate BAT thermogenesis, which often simultaneously recruits other thermogenic mechanisms. For instance, much of what is known about human BAT function comes from cold-stimulated conditions, where thermogenesis and substrate clearance is primarily driven by shivering skeletal muscles ^9–12^, even under very mild conditions ^13^. Alternatively, the recent repurposing of the β_3_-adrenergic receptor (β_3_-AR) agonist mirabegron, a medication used to treat overactive bladder (OAB), has provided the opportunity to isolate the metabolic effects of targeting BAT more specifically given its high affinity for the human β_3_-AR compared to the β_2_- or β_1_-AR (105-fold greater affinity for β_3_-AR than β_1_-AR, and 33-fold stronger than β_2_-AR).

The initial investigations using mirabegron to stimulate BAT revealed two important trends ^14–18^. First, stimulating BAT thermogenesis required a dosage that was four-times higher than the maximum approved dosage (approved for 50 mg daily) ^19^. Only when mirabegron was ingested at the maximal allowable dose (200 mg) in lean healthy men did whole-body energy expenditure and BAT glucose uptake increase ^17, 19^. Furthermore, only the high dose of mirabegron increased BAT oxidative metabolism compared to room temperature, levels that remained significantly lower than what is observed during mild cold exposure ^19^. Secondly, such a high dose of mirabegron led to nonselective β-AR activation, resulting in an increase in cardiovascular responses (β_1_-AR- mediated) and white adipose tissue (WAT) lipolysis (primarily β_2_-AR-mediated). Indeed, both heart rate and systolic blood pressure increased when given the maximal allowable dose of mirabegron, consistent with the secondary effects reported in the early mirabegron clinical trials for the treatment of OAB ^20–22^. In humans, the heart expresses mainly the β_1_-AR ^23, 24^ while brown adipocytes express mainly the β_2_-AR and β_1_-AR and very low levels of β_3_-AR ^19, 25, 26^. This explains why, despite its high affinity for the β_3_-AR, mirabegron may still bind to the β_2_-AR and β_1_-AR to stimulate thermogenesis. We also recently showed that silencing the gene encoding the β_2_-AR (*ADRB2*) *in vitro*, resulted in a compensatory increase in the expression of the gene encoding the β_3_-AR (*ADRB3*) ^19^. Similarly, others have shown that silencing the gene encoding for the β_1_-AR (*ADRB1*) significantly increases brown/beige adipocyte oxygen consumption, despite decreasing *UCP1* expression^27^. In light of these findings, the primary aim of this study was to examine whether administration of the maximal allowable dose of mirabegron when combined with a cardio-selective β_1_-AR antagonist, bisoprolol, could increase the sensitivity of the β_3_-AR and β_2_- AR in WAT and BAT while suppressing the unwanted cardiovascular effects that present with the administration of high doses of mirabegron. Thus, we hypothesized that administration of mirabegron with a β_1_-AR receptor antagonist could block cardiovascular effects while increasing BAT thermogenesis above levels observed when mirabegron is administered alone.

## RESULTS

### The Beneficial Metabolic Responses Induced by Mirabegron Are Attenuated by Bisoprolol

The main purpose of this study was to compare the stimulation of BAT metabolism using mirabegron without or with a β_1_-AR antagonist in 8 lean healthy individuals (Table 1, Figure 1). We hypothesized that the acute oral administration of the maximal allowable dose of mirabegron (200 mg) with the β_1_-AR antagonist, bisoprolol (10 mg), would result in an increase in BAT oxidative metabolism and whole-body energy expenditure without the adverse cardiovascular events observed when a high dose of mirabegron is administered alone. The primary outcome measures of this clinical study as entered in clinicaltrials.gov (NCT04823442) included the change in activation of BAT (oxidative metabolism and blood flow) and BAT net glucose uptake. Secondary endpoints can be found at https://clinicaltrials.gov/ct2/show/NCT04823442.

**Figure 1.**
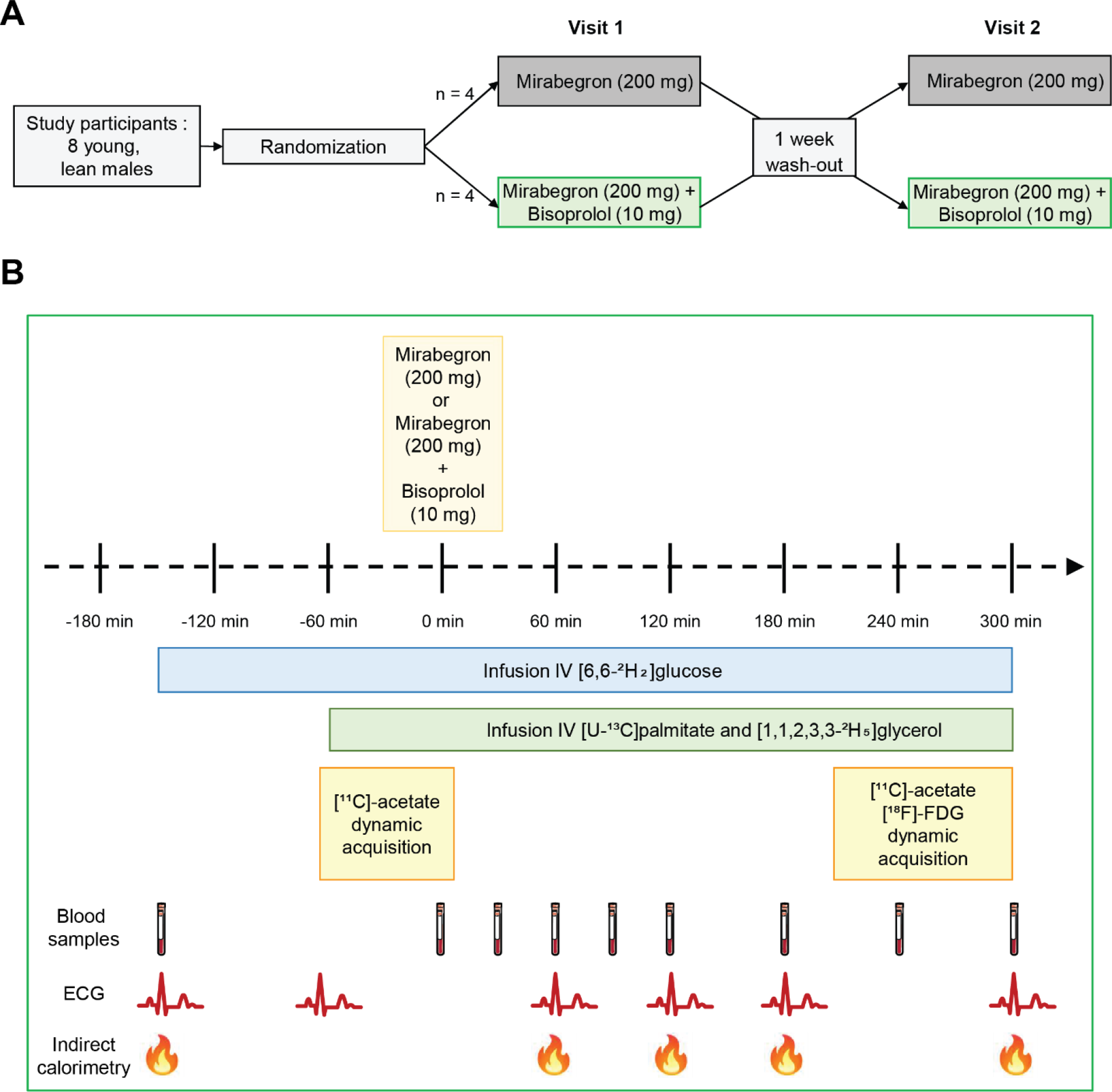
Study protocol. (A) Randomized crossover study (n=8) where participants received mirabegron (200 mg) without or with bisoprolol (10 mg), with a one-week wash-out period between visits. (B) Metabolic protocol for both study visits.

**Table 1.** Participant characteristics

| **Participant Characteristics** | **Healthy men** |
| --- | --- |
| **N** | 8 |
| **Age (years)** | $27 \pm 3$ |
| **Height (cm)** | $181.5 \pm 3.9$ |
| **Mass (kg)** | $74.1 \pm 5.8$ |
| **BMI (kg/m<sup>2</sup>)** | $22.5 \pm 1.7$ |
| **Fat mass (kg)** | $19.6 \pm 5.3$ |
| **Lean mass (kg)** | $53.5 \pm 6.1$ |
| **Waist circumference (cm)** | $80.8 \pm 6.9$ |
| **Hip circumference (cm)** | $98.4 \pm 4.0$ |
| **Body fat (%)** | $26.6 \pm 5.3$ |

BMI, Body Mass Index
Data expressed as mean ± SD

Using liquid chromatography-tandem mass spectrometry we first examined the plasma pharmacokinetics of mirabegron without (M) or with bisoprolol (MB) in the participants. The study was designed to ensure that the time of maximal plasma mirabegron concentrations coincided with our dynamic list-mode PET acquisition which was 240 minutes after oral administration of the mirabegron without or with bisoprolol (Table 2 and Figure 2A; Median: M: 180 min; MB: 240 min). Compared to baseline measures, oral administration of either mirabegron alone or in combination with bisoprolol increased resting energy expenditure by 27% (+18.2 ± 6.7 kcal/h; P < 0.0001) and 11% (+7.4 ± 8.1 kcal/h; P = 0.0268), respectively (Figure 2B; Table 2). However, the resting energy expenditure was significantly lower when mirabegron was co-ingested with bisoprolol compared to when it was ingested alone (Figure 2 C; P = 0.0402).

**Figure 2.**
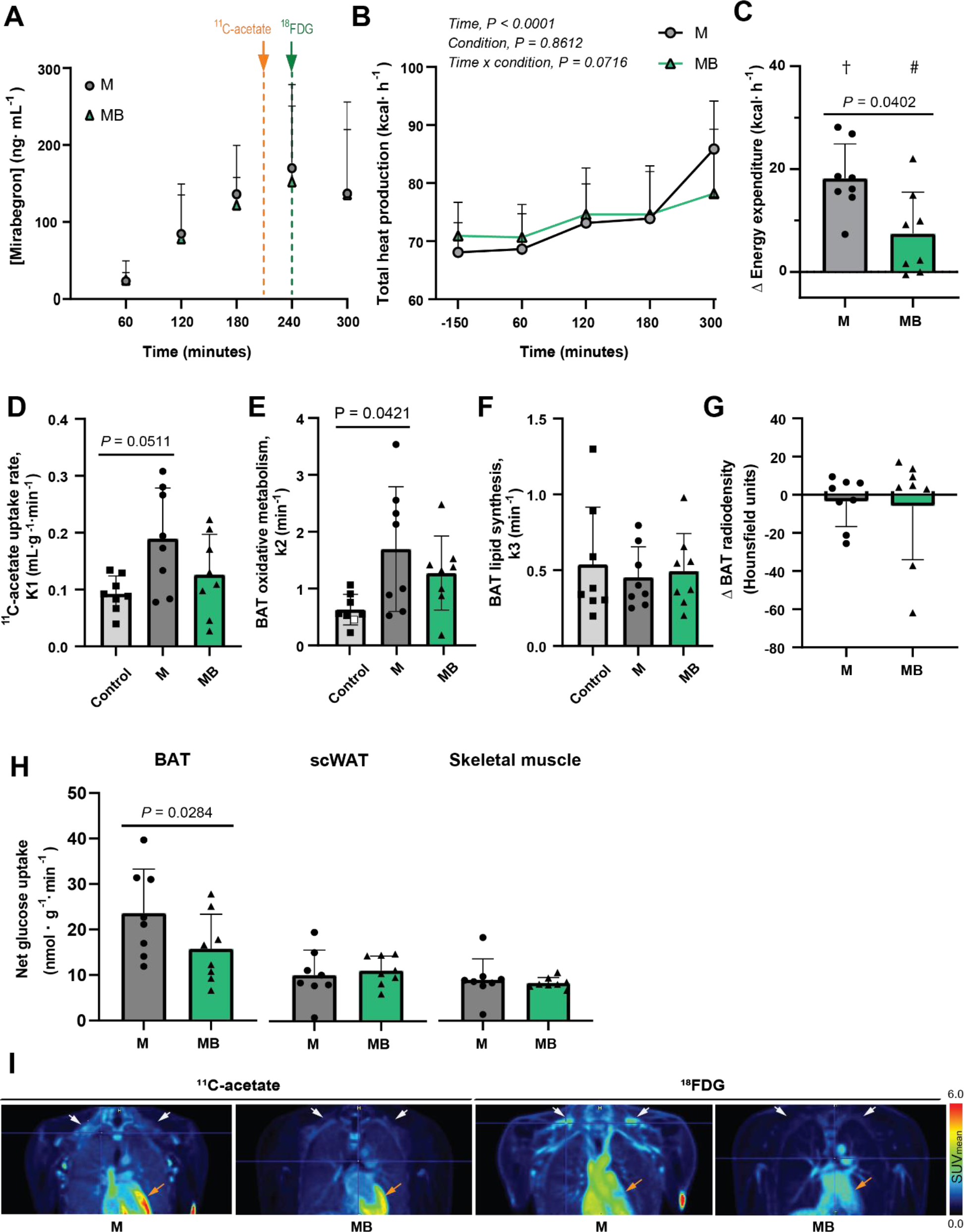
Mirabegron increases BAT oxidative metabolism and glucose uptake but not in combination with bisoprolol. (A) Circulating mirabegron levels following oral administration of mirabegron (M, 200 mg) without or with bisoprolol (MB, 10 mg) in humans. [^11^C]acetate (orange arrow) and 2-deoxy-2- [^18^F]-fluoro-D-glucose ([^18^F]FDG, green arrow) radiotracers were injected 210 and 240 min after oral administration of mirabegron or mirabegron + bisoprolol, respectively, followed by a dynamic list-mode PET acquisition (n = 8). (B and C) Whole body energy expenditure over time (B) and change from basal levels (C) in response to oral administration of mirabegron (M, 200 mg) without or with bisoprolol (MB,10 mg) (n = 8). The effect of intervention was determined using two-way ANOVA for repeated-measures with Bonferroni post hoc test. # *P* ≤ 0.05, ǂ *P* ≤ 0.0001. (D-F) [^11^C]acetate uptake rate (D, *K1*), oxidative metabolism (E, *k2*) and lipid synthesis (F, *k3*) of supraclavicular BAT at baseline (Control) and in response to oral administration of mirabegron (M, 200 mg) without or with bisoprolol (MB, 10 mg). Outliers were identify using the Interquartile Range Method. One outlier (white square) was detected and replaced by the mean of this group and a previous study^42^. (G) Change in supraclavicular BAT radiodensity from basal levels in responses to oral administration of mirabegron (M, 200 mg) without or with bisoprolol (MB, 10 mg) (n = 8). (H) Supraclavicular BAT, WAT, and skeletal muscle net glucose uptake in response to oral administration of mirabegron (M, 200 mg) without or with bisoprolol (MB, 10 mg) (n = 8). Skeletal muscle glucose uptake represents mean uptake of regions of interest drawn from *m. pectoralis major, m. trapezius, m. deltoideus, m. sternocleidomastoid, m. levator scapulae, m. latissimus dorsi,* and *m. erector spinae*. (I) Cervico-thoracic PET images of [^11^C]acetate (two right images) and [^18^F]FDG uptake in response to oral administration of mirabegron (M, 200 mg) without or with bisoprolol (MB, 10 mg). PET images of [^11^C]acetate taken at 210 min and [^18^F]FDG taken at 240 min in the same participant are depicted in response to mirabegron without or with bisoprolol. White arrows represent supraclavicular BAT depot and orange arrows represent heart. Data presented as mean ± SD. A One-way ANOVA for repeated measures with Bonferroni *post hoc* test was used to determine statistically significant differences between the basal and the two treatment conditions. Difference between mirabegron without or with bisoprolol determined using paired-sample t test.

**Table 2.**
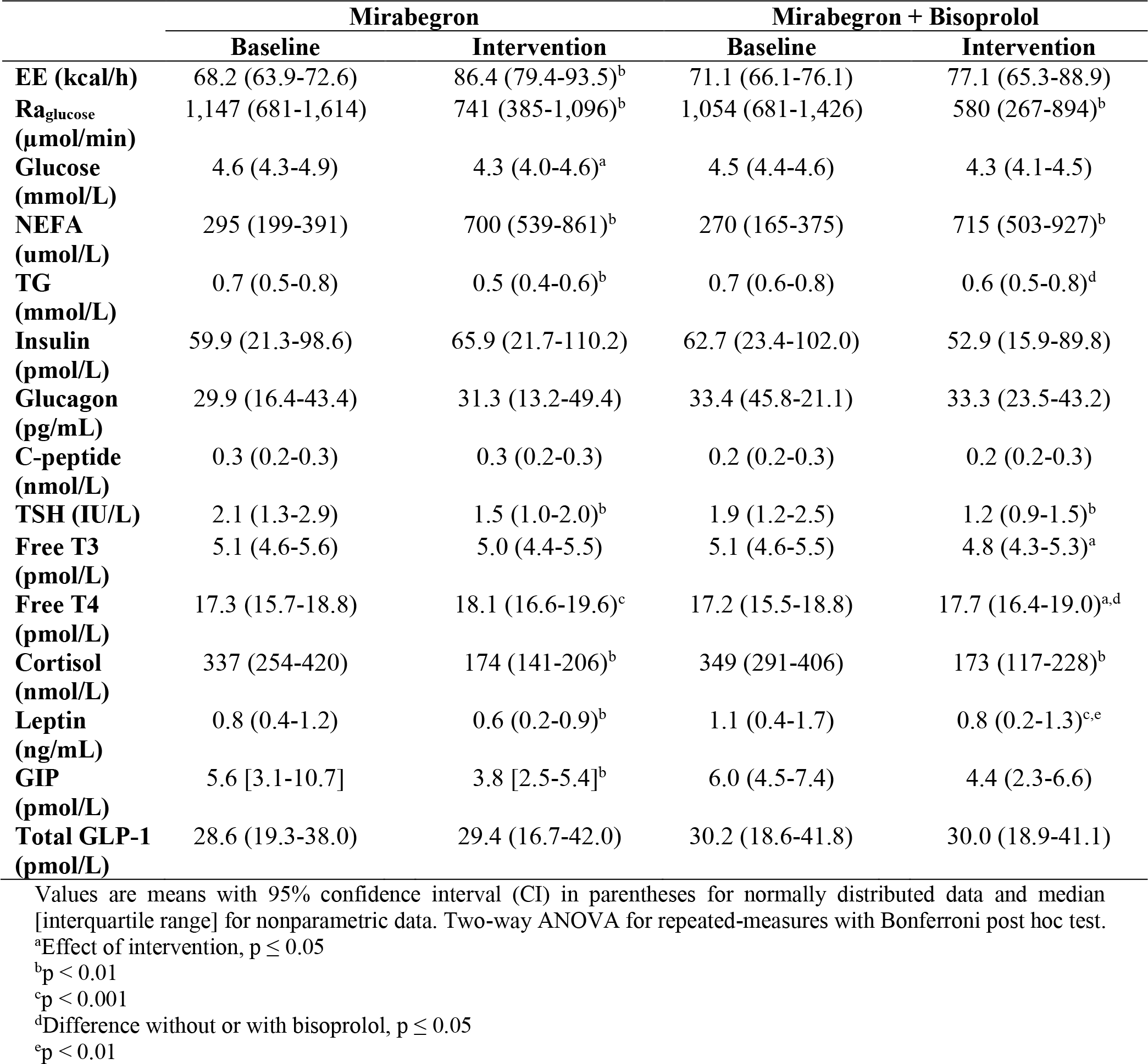
Hormone and metabolite concentrations at baseline or with the ingestion of 200 mg mirabegron without or with 10 mg of bisoprolol

|  | Mirabegron |  | Mirabegron + Bisoprolol |  |
| --- | --- | --- | --- | --- |
|  | Baseline | Intervention | Baseline | Intervention |
| **EE (kcal/h)** | 68.2 (63.9-72.6) | 86.4 (79.4-93.5)<sup>b</sup> | 71.1 (66.1-76.1) | 77.1 (65.3-88.9) |
| **R<sub>a</sub>glucose (μmol/min)** | 1,147 (681-1,614) | 741 (385-1,096)<sup>b</sup> | 1,054 (681-1,426) | 580 (267-894)<sup>b</sup> |
| **Glucose (mmol/L)** | 4.6 (4.3-4.9) | 4.3 (4.0-4.6)<sup>a</sup> | 4.5 (4.4-4.6) | 4.3 (4.1-4.5) |
| **NEFA (μmol/L)** | 295 (199-391) | 700 (539-861)<sup>b</sup> | 270 (165-375) | 715 (503-927)<sup>b</sup> |
| **TG (mmol/L)** | 0.7 (0.5-0.8) | 0.5 (0.4-0.6)<sup>b</sup> | 0.7 (0.6-0.8) | 0.6 (0.5-0.8)<sup>d</sup> |
| **Insulin (pmol/L)** | 59.9 (21.3-98.6) | 65.9 (21.7-110.2) | 62.7 (23.4-102.0) | 52.9 (15.9-89.8) |
| **Glucagon (pg/mL)** | 29.9 (16.4-43.4) | 31.3 (13.2-49.4) | 33.4 (45.8-21.1) | 33.3 (23.5-43.2) |
| **C-peptide (nmol/L)** | 0.3 (0.2-0.3) | 0.3 (0.2-0.3) | 0.2 (0.2-0.3) | 0.2 (0.2-0.3) |
| **TSH (IU/L)** | 2.1 (1.3-2.9) | 1.5 (1.0-2.0)<sup>b</sup> | 1.9 (1.2-2.5) | 1.2 (0.9-1.5)<sup>b</sup> |
| **Free T3 (pmol/L)** | 5.1 (4.6-5.6) | 5.0 (4.4-5.5) | 5.1 (4.6-5.5) | 4.8 (4.3-5.3)<sup>a</sup> |
| **Free T4 (pmol/L)** | 17.3 (15.7-18.8) | 18.1 (16.6-19.6)<sup>c</sup> | 17.2 (15.5-18.8) | 17.7 (16.4-19.0)<sup>a,d</sup> |
| **Cortisol (nmol/L)** | 337 (254-420) | 174 (141-206)<sup>b</sup> | 349 (291-406) | 173 (117-228)<sup>b</sup> |
| **Leptin (ng/mL)** | 0.8 (0.4-1.2) | 0.6 (0.2-0.9)<sup>b</sup> | 1.1 (0.4-1.7) | 0.8 (0.2-1.3)<sup>c,e</sup> |
| **GIP (pmol/L)** | 5.6 [3.1-10.7] | 3.8 [2.5-5.4]<sup>b</sup> | 6.0 (4.5-7.4) | 4.4 (2.3-6.6) |
| **Total GLP-1 (pmol/L)** | 28.6 (19.3-38.0) | 29.4 (16.7-42.0) | 30.2 (18.6-41.8) | 30.0 (18.9-41.1) |

Values are means with 95% confidence interval (CI) in parentheses for normally distributed data and median [interquartile range] for nonparametric data. Two-way ANOVA for repeated-measures with Bonferroni post hoc test.
<sup>a</sup>Effect of intervention, $p \leq 0.05$
<sup>b</sup> $p < 0.01$
<sup>c</sup> $p < 0.001$
<sup>d</sup>Difference without or with bisoprolol, $p \leq 0.05$
<sup>e</sup> $p < 0.01$

A multi-tracer approach with [^11^C]acetate and the [^18^F]FDG was used to evaluate the rate of BAT oxidative metabolism and tissue-specific glucose uptake. A four-compartment, two-tissue pharmacokinetic model for [^11^C]acetate in BAT^28^ was applied to derive the rates of [^11^C]acetate uptake (*K*_1_, mL·g^-1^·min^-1^), a marker of tissue perfusion, oxidative metabolism (*k*_2_, min^-1^) and lipid synthesis (*k*_3_, min^-1^). The rate of BAT [^11^C]acetate uptake increased with mirabegron alone (Figure 2D; P = 0.0511), which was accompanied by a 2.7-fold increase in BAT oxidative metabolism (Figure 2E; from 0.636 ± 0.251 min^-1^ at room temperature to 1.629 ± 1.097 min^-1^ with mirabegron alone, P = 0.0421). Nevertheless, BAT oxidative metabolism was not significantly different when the mirabegron was co-ingested with the bisoprolol compared to control room temperature conditions (Figure 2E; from 0.636 ± 0.251 min^-1^ at room temperature to 1.273 ± 0.651 min^-1^). We found that mirabegron without or with bisoprolol did not elicit a significant increase in BAT intracellular lipid synthesis (Figure 2F), nor elicit a difference in the change in CT-derived tissue radiodensity, which represents a marker of changes in intracellular triglyceride content (Figure 2G). Using the Patlak linearization model, we showed that the high dose of mirabegron increased the rate of BAT glucose uptake, but was significantly lower when combined with bisoprolol (24 ± 10 nmol‧g^-1^‧min^-1^ with mirabegron alone *vs.* 16 ± 8 nmol‧g^-1^‧min^-1^ with mirabegron and bisoprolol; P = 0.0284). Although the rate of BAT glucose uptake was lower when mirabegron was given with bisoprolol, it remained significantly greater than what has previously been reported under room temperature conditions [9 ± 4 nmol‧g^-1^‧min^-1^, N = 27; P *=* 0.0018^29^]. We did not observe any differences in net glucose uptake in subcutaneous WAT (scWAT; 12 ± 7 nmol‧g^-1^‧min^-1^ with mirabegron alone *vs.* 13 ± 4 nmol‧g^-1^‧min^-1^ with mirabegron combined with bisoprolol) or skeletal muscles (from 11 ± 6 nmol‧g^-1^‧min^-1^ with mirabegron alone *vs.* 10 ± 1 nmol‧g^-1^‧min^-1^ with mirabegron combined with bisoprolol) between the two conditions (Figure 2H).

### Bisoprolol Suppressed Cardiovascular Secondary Effects of Mirabegron

It has previously been demonstrated that administration of high doses of mirabegron leads to unwanted cardiovascular effects, likely mediated through a cross-activation of the β_1_-AR ^17, 18, 30, 31^. These undesirable effects are what prompted us to assess cardiovascular outcomes when combining mirabegron with the cardioselective β_1_-AR antagonist, bisoprolol ^32^. Mirabegron alone increased systolic blood pressure by 5 ± 5 mmHg above baseline levels. In contrast, when mirabegron was combined with bisoprolol, systolic blood pressure decreased below baseline levels by 6 ± 10 mmHg (Figure 3A, P = 0.0252). Diastolic blood pressure decreased by 3 ± 6 mmHg and 4 ± 19 mmHg below pre-treatment levels with mirabegron alone or with bisoprolol, respectively, with no significant difference between the two conditions (Figure 3B). Mirabegron given alone increased heart rate by 6 ± 7 beats‧min^-1^ (P = 0.0565) above pre-treatment levels, whereas mirabegron administered with bisoprolol decreased heart rate by 5 ± 6 beats‧min^-1^ (P = 0.1302) below the pre-treatment levels (Figure 3C, P = 0.0081). To examine the effects of mirabegron with or without bisoprolol on the myocardial metabolic demands, we calculated the rate pressure product (RPP). While RPP increased significantly by 972 ± 927 mmHg‧bpm above pre-treatment levels (P = 0.0333) when given mirabegron alone, it decreased by 1548 ± 1158 mmHg‧bpm compared to pre-treatment levels when mirabegron was administered with bisoprolol (P = 0.0544) which was significantly different between the two conditions (Figure 3D, P = 0.0005).

**Figure 3.**
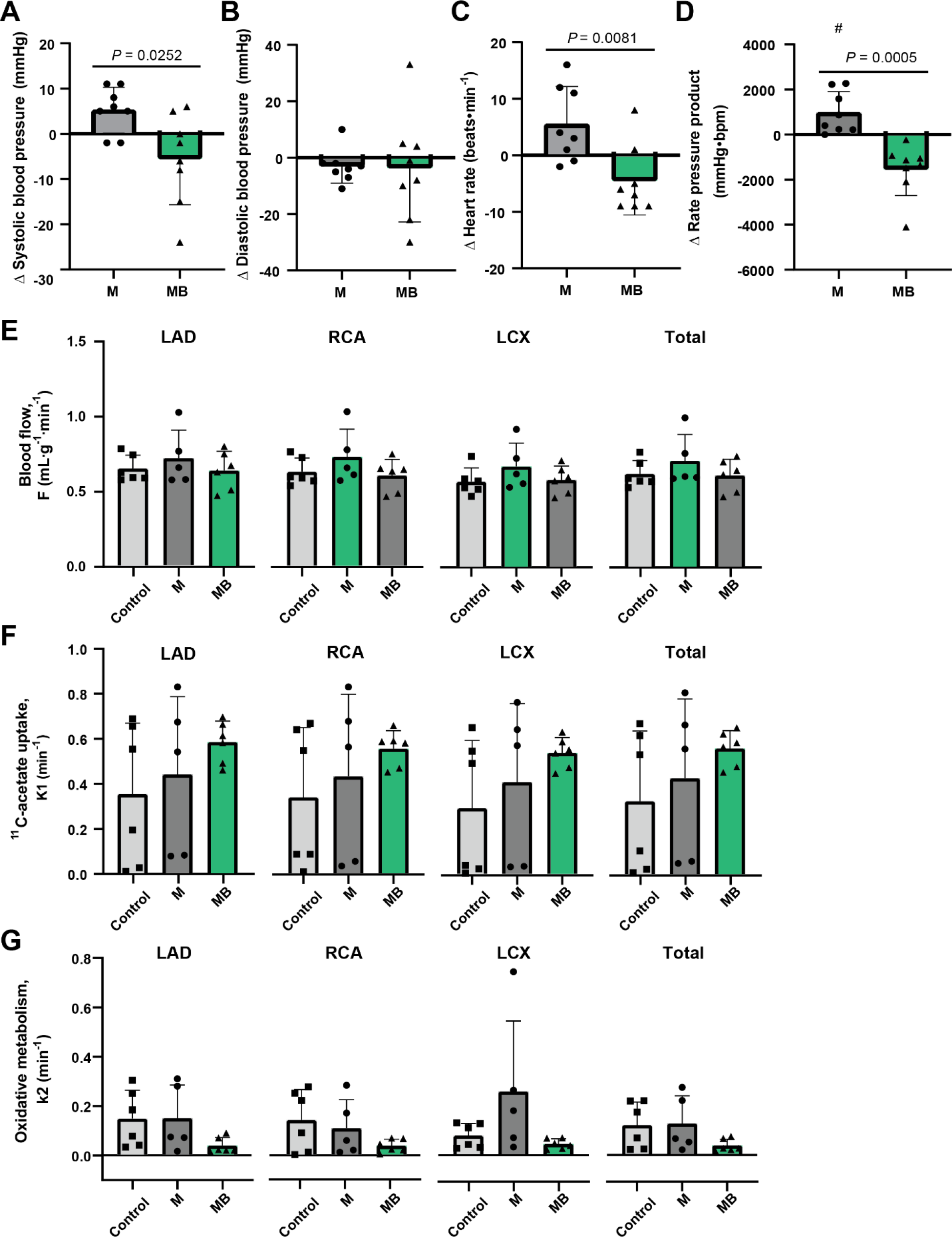
Mirabegron with bisoprolol suppressed the cardiovascular side effects of mirabegron alone. (A-C) Change in systolic blood pressure (A), diastolic blood pressure (B), and heart rate (C) from basal levels in response to oral administration of mirabegron (M, 200 mg) without or with bisoprolol (MB,10 mg) (n = 8). (D) Change in rate pressure product from basal levels in response to oral administration of mirabegron (M, 200 mg) without or with bisoprolol (MB, 10 mg) (n = 8). The effect of intervention was determined using two-way ANOVA for repeated-measures with Bonferroni post hoc test. # *P* ≤ 0.05. (E) Blood flow (F) obtained from the [^11^C]acetate analysis of three segments (LAD, RCA and LCX) of the heart and total heart (Global) at baseline (Control) and in response to oral administration of mirabegron (M, 200 mg) without or with bisoprolol (MB, 10 mg) (n = 5-6). (F) [^11^C]acetate uptake (*K1*) of three segments (LAD, RCA and LCX) of the heart and total heart (Global) at baseline (Control) and in response to oral administration of mirabegron (M, 200 mg) without or with bisoprolol (MB, 10 mg) (n = 5-6). (G) Oxidative metabolism (*k2*)obtained from the [^11^C]acetate analysis of three segments (LAD, RCA and LCX) of the heart and total heart (Global) at baseline (Control) and in response to oral administration of mirabegron (M, 200 mg) without or with bisoprolol (MB, 10 mg) (n = 5-6). LAD: Left anterior descending artery; LCX: Left circumflex artery; RCA: Right coronary artery. Data presented as mean ± SD. Difference between mirabegron without or with bisoprolol determined using paired-sample t test

As result of the large field of view of the PET scanner (26 cm), it was possible for the first time to simultaneously quantify BAT thermogenesis and myocardial oxygen consumption in humans. Myocardial blood flow (F), tracer uptake (*K*_1_) and oxidative metabolism (*k*_2_) were assessed by [^11^C]acetate PET in three segments of the heart vascularized by the left anterior descending artery (LAD), left circumflex artery (LCX), and right coronary artery (RCA) as well as globally (total), pre- and post-treatment (Figure 3E, 3F, and 3G). In order to do this, we analyzed PET dynamic images of the heart with the PMOD cardiac PET modeling tool, PCARDP (PMOD Technologies, version 3.7). However, since the PET scanner has a limited field of view and participants varied in height, we prioritized PET image acquisition of supraclavicular BAT at the expense of having the complete image of the heart. Therefore, we have a limited number of PET images for the analysis of the heart, which did not allow a complete statistical analysis. However, based on the [^11^C]acetate data, myocardial blood flow did not appear to be affected by mirabegron, whether administered alone or with bisoprolol, when compared to pre-treatment conditions, in all the regions of the heart (Figure 3E). The [^11^C]acetate uptake in the heart increased when given mirabegron compared to pre-treatment levels and increased further in combination with the bisoprolol (Figure 3F). The oxidative metabolism of the three segments of the myocardium as well as global myocardium oxidative metabolism were more elevated in the pre-treatment condition and when mirabegron was given alone compared to the administration of mirabegron with bisoprolol. The reduction in oxidative metabolism when mirabegron was administered in combination with bisoprolol is consistent with the reduction in RPP, which is commonly used as an index of myocardial oxygen consumption ^33, 34^.

### Mirabegron Without or With Bisoprolol Did Not Alter White Adipose Tissue Lipolysis

Since the high dose of mirabegron leads to nonselective β-AR activation, we assessed white adipose tissue (WAT) lipolysis which is primarily driven by β_2_/β_1_-AR signaling. To examine this response we used continuous infusion of [U-^13^C]palmitate and [1,1,2,3,3-_2_H^5^]-glycerol to determine the rate of systemic appearance of NEFA and glycerol (indicator of whole-body WAT lipolysis) (Figure 4A-4D). The systemic appearance rate (R_a_) of glycerol (Ra_glycerol_) and NEFA (Ra_NEFA_) did not increase significantly (Figure 4A-4D). However, compared to the baseline period, the rate of fatty acid oxidation increased significantly both when mirabegron was given alone (P = 0.0021) and when given in combination with bisoprolol (P = 0.0038) (Figure 4F), however there was no significant difference between the two conditions. In steady-state conditions, the difference between the total fatty acids released upon intracellular hydrolysis of triglycerides (3 X Ra_glycerol_) and the rate of total fatty acid oxidation (calculated from indirect calorimetry) indicate the whole-body rate of fatty acid re-esterification, as this is primarily its alternative metabolic fate. Since the increase in both the rate of whole-body lipolysis and fatty acid oxidation were not different whether mirabegron was given alone or with bisoprolol, this suggests that there was no difference in fatty acid re-esterification between the two groups (Figure 4I). Indeed, there was no change in the re-esterification of the fatty acids at the primary site of lipolysis (intracellular cycling) or following their transit in circulation where they can be taken up by the liver to be stored in intracellular lipid droplets or incorporated into VLDL (extracellular cycling) (Figure 4I).

**Figure 4.**
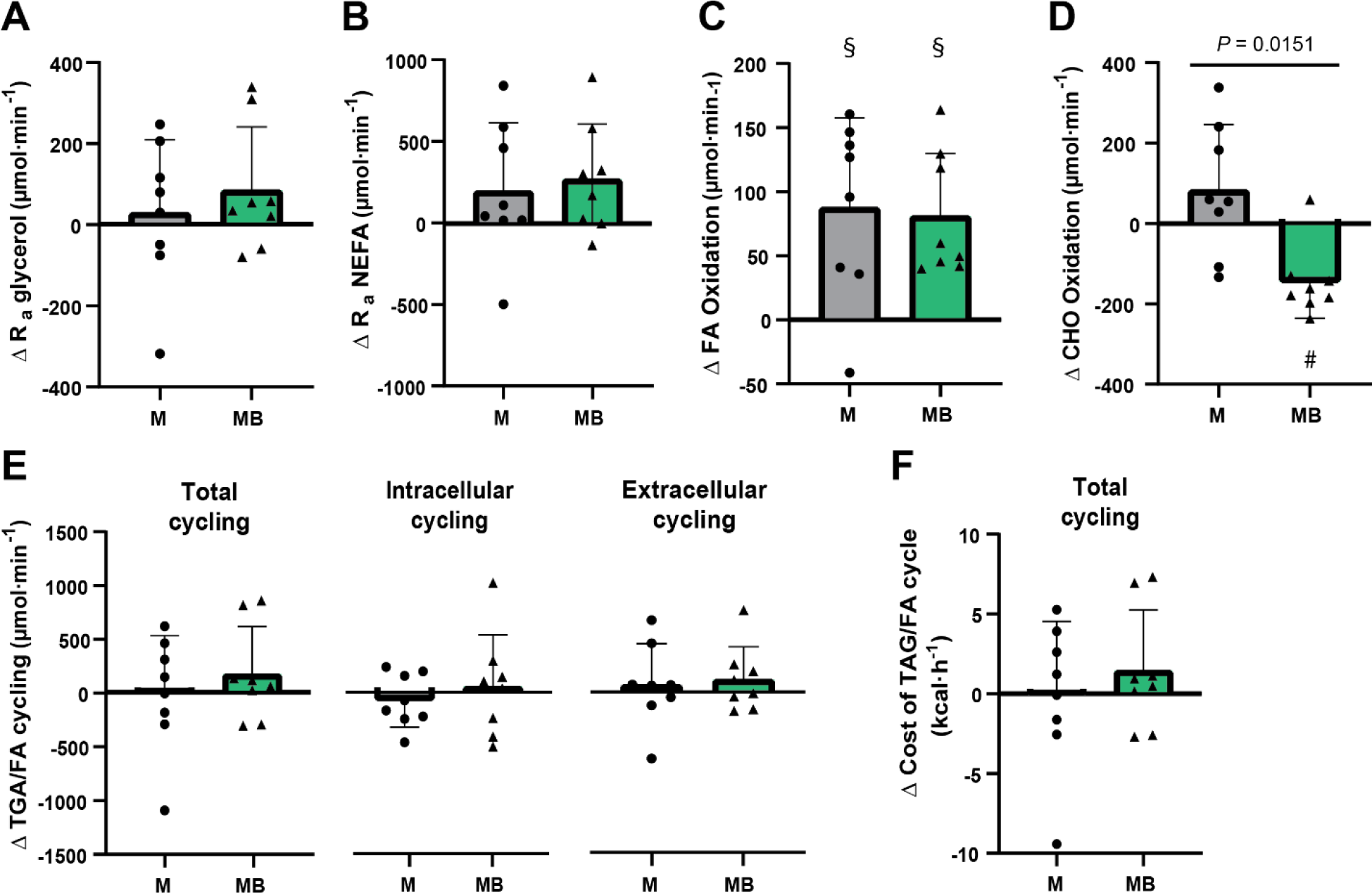
Mirabegron alone or in combination with bisoprolol did not impact the TAG/NEFA cycling. (A and B) Change in systemic rate of appearance of glycerol over time (A) and change from basal levels (B) in response to oral administration of mirabegron (M, 200 mg) without or with bisoprolol (MB,10 mg) as index of whole-body lipolysis (n = 8). (C and D) Change in systemic rate of appearance of NEFA over time (C) and change from basal levels (D) in response to oral administration of mirabegron (M, 200 mg) without or with bisoprolol (MB, 10 mg) (n = 8). (E and F) Change in whole-body fatty acid oxidation over time (E) and change from basal levels (F) in response to of mirabegron (M, 200 mg) without or with bisoprolol (MB,10 mg) determined by indirect calorimetry (n = 8). The effect of the intervention was determined using two-way ANOVA for repeated-measures with Bonferroni post hoc test. # *P* ≤ 0.05, § *P* ≤ 0.001. (G and H) Change in whole-body glucose utilization over time (G) and change from basal levels (H) in response to mirabegron (M, 200 mg) without or with bisoprolol (MB,10 mg) determined by indirect calorimetry (n = 8). The effect of intervention was determined using two-way ANOVA for repeated-measures with Bonferroni post hoc test. # *P* ≤ 0.05, § *P* ≤ 0.001 (I) Change in total, intracellular, and extracellular triacylglycerol/fatty acid (TAG/FA) cycling from basal levels in response to of mirabegron (M, 200 mg) without or with bisoprolol (MB,10 mg) (n = 8). (J) Change in the energetic cost of total TAG/FA cycle (n = 8). Data presented as mean ± SD. Difference between mirabegron without or with bisoprolol determined using paired-sample t test.

## DISCUSSION

The ingestion of the β_3_-AR agonist mirabegron at the maximal allowable dose (200 mg) increases BAT thermogenesis but also leads to an adverse overstimulation of cardiovascular responses, likely by stimulating all three β-AR subtypes ^17, 18, 30, 31^. Here we show that combining the administration of mirabegron with a cardioselective β_1_-AR antagonist (bisoprolol; 15-fold more selective for β_1_-AR vs β_2_-AR and 31-fold more selective for β_1_-AR vs β_3_-AR^35^), to suppress these cardiovascular responses, did not increase BAT oxidative metabolism, and in fact reduced BAT blood flow. This was also accompanied by a reduction in the rate of BAT glucose uptake. By design, bisoprolol did indeed inhibit the mirabegron-stimulated increase in systolic blood pressure and heart rate. It is unclear whether the attenuation of systemic and BAT blood flow induced by the relatively large dose of bisoprolol used may have contributed to the impaired BAT thermogenesis.

Previously, we showed that oral administration of the maximal allowable dose of mirabegron (200 mg) increases BAT oxidative metabolism on average 2.1-fold above basal conditions. This was also accompanied by a 17% increase in whole-body energy expenditure ^19^. The aim of the present study was, first, to examine whether giving this same dose of mirabegron in combination with a cardioselective β_1_-AR antagonist could increase BAT oxidative metabolism while blunting the cardiovascular responses. We were also interested in determining whether inhibiting the β_1_-AR could increase the sensitivity of the β_2_- and β_3_-AR. We previously showed that silencing the *ADRB2 in vitro* resulted in a compensatory increase in the expression of *ADRB3* ^19^. We therefore hypothesized that inhibiting the β_1_-AR would not only prevent the undesirable cardiovascular responses, but also increase the stimulation of the β_2_-AR and β_3_-AR, thereby allowing mirabegron to be administered at a lower or similar dose while eliciting similar or greater thermogenic effects. Indeed, whether mirabegron was administered alone or in combination with bisoprolol, energy expenditure increased above baseline levels (by 27% with mirabegron alone and 11% with mirabegron and bisoprolol). Similar to our previous investigation, BAT oxidative metabolism and BAT blood flow increased with the high dose of mirabegron administration. However, in contrast to our hypothesis, this effect was blunted when mirabegron was administered with bisoprolol. The exact cause of this reduction in BAT oxidative metabolism remains unclear. One explanation could be that β_1_-AR inhibition, rather than potentiating the effect of mirabegron, directly inhibited BAT oxidative metabolism. This would certainly be consistent with a previous *in vitro* study, which showed that silencing *ADRB1* in immortalized human brown adipocytes significantly attenuated *UCP1* induction in response to isoproterenol stimulation ^36^. However, we and others have shown that silencing *ADRB1* in human primary brown adipocytes has no effect on cellular respiration, whether stimulated with norepinephrine ^19^ or not ^37^. Alternatively, a more plausible explanation is that the reduction in BAT blood flow when mirabegron was given with bisoprolol restricted the ability for BAT to increase oxidative metabolism. Although adrenergic stimulation can increase BAT blood flow, even in the absence of thermogenesis in mice ^38^ and humans ^39^, the suppression of BAT blood flow in male Wistar rats has been shown to significantly reduce the norepinephrine-stimulated increase in BAT temperature ^40^. Evidence from the present study suggest this may also be the case in humans.

Whether this reduction in BAT blood flow is a direct effect of β_1_-AR-antagonism on BAT vasomotor tone or an indirect result of a possible decrease in cardiac output remains unclear. The rate of glucose uptake was also reduced when bisoprolol was given with mirabegron, which may have been partially mediated by the reduction in BAT blood flow. It should be noted that the rate of glucose uptake was still significantly greater than what has been reported under unstimulated room temperature conditions (16 ± 8 nmol·g^-1^·min^-1^ with mirabegron + bisoprolol vs 9 ± 4 nmol·g^-1^·min^-1^ at room temperature; ^41^) or when given a therapeutic dose (50 mg) of mirabegron (9 ± 5 nmol·g^-1^·min^-1^; ^19^) One noted secondary effect of mirabegron administration is the increase in systolic blood pressure and heart rate when it is administered at the maximal allowable dose. Consistent with previous findings, the administration of the maximal allowable dose of mirabegron increased systolic blood pressure and heart rate by 5 ± 5 mmHg and 6 ± 7 beats‧min^-1^ above pre-treatment levels, respectively. In contrast, when mirabegron was given with bisoprolol, heart rate and rate pressure product (RPP), an indicator of myocardial oxygen consumption, decreased below baseline levels. The reduction in RPP agrees with the results obtained by [^11^C]acetate PET imaging of the myocardium performed in a sub-group of participants, which showed a global reduction in myocardial oxidative metabolism with bisoprolol. All these results were expected given the cardioselective nature of the β_1_-AR antagonist used. The question remains whether this decrease in myocardial metabolism and cardiovascular responses contributed to the reduction in BAT thermogenesis.

In our previous investigation, we demonstrated that oral administration of mirabegron, not only stimulated BAT thermogenesis, but also increased the systemic rate of appearance of NEFA and glycerol, an indicator of whole-body WAT lipolysis, by 115% and 126%, respectively ^19^. The increased WAT lipolysis far exceeded the rate of whole-body fatty acid oxidation. This suggested that a significant proportion of the fatty acids released following the hydrolysis of intracellular triglycerides were likely re-esterified *in situ*. We estimated that this energetically-costly triglyceride hydrolysis-fatty acid re-esterification cycle, commonly referred to as the triglyceride- fatty acid (TAG-FA) cycle or glycerolipid-free fatty acid (GL-FFA) cycle, accounted for 55% of the mirabegron-stimulated increase in energy expenditure. In the present study, we did not observe a significant increase in the systemic appearance of glycerol and NEFA. Consequently, we did not observe significant changes in the TAG-FA cycle and therefore it was likely a minor contributor to the increase in energy expenditure in the present study, accounting for 5.6% of the increase in energy expenditure following mirabegron administration alone, and 19.6% when mirabegron was given with bisoprolol. The slightly lower whole-body energy expenditure with mirabegron and bisoprolol administration was also accompanied by a decrease in carbohydrate utilization, which might reflect the transient decrease in the systemic rate of appearance of glucose.

## LIMITATIONS OF THE STUDY

There are a few limitations to consider in the interpretation of our findings. First, this study included a small number of young, healthy men. Only male participants were recruited, as the dose given to stimulate BAT was 2-fold greater than the maximal allowable limit permitted for women ^31^. In women, the ingestion of mirabegron is associated with a dose-dependent prolongation of the QTc interval, thus restricting the maximum allowable dose to 100 mg. Further work is needed to examine whether there are sex-dependent differences in mirabegron-mediated activation of BAT metabolism. Second, the high dose of bisoprolol used significantly decreased heart rate and the systolic blood pressure below baseline levels, which could have contributed to the observed reduction in BAT blood flow and could have limited the ability to stimulate BAT thermogenesis. The significant reduction in cardiovascular responses was somewhat limiting as there were concerns with inducing significant levels of bradycardia. Indeed, of the 9 participants recruited one participant with a particularly low resting heart rate (45 bpm) was precluded from completing the mirabegron + bisoprolol condition as there were concerns about further decreasing his heart rate. Finally, the PET/CT scanner used in the present study has a relatively large field of view (26 cm), which for the first time, allowed for the simultaneous quantification of BAT thermogenesis and myocardial oxygen consumption in humans. However, in some of our taller participants, it was not possible to image both the BAT and heart and therefore we prioritized the acquisition of supraclavicular BAT. This resulted in an incomplete dataset of the [^11^C]acetate PET pharmacokinetic analysis of the myocardium.

## CONCLUSION

In conclusion, as demonstrated in our previous study, a single maximal allowable dose of mirabegron elicited a significant increase in human BAT thermogenesis and whole-body energy expenditure. In contrast to our initial hypothesis, the addition of bisoprolol did reduce mirabegron-stimulated BAT oxidative metabolism. Whether the attenuation in BAT blood flow induced by the large dose of bisoprolol led to this reduction in mirabegron-stimulated BAT thermogenesis remains to be determined. Further work is needed to determine the optimal pharmacological approach to activate BAT thermogenesis in humans.

## Data Availability

All data produced in the present study are available upon reasonable request to the authors.

## Acknowledgements

The authors would like to thank the participants of this study for their commitment and collaboration. The authors thank Caroll-Lynn Thibodeau, Maude Gérard, Myriam Flipot, Éric Lavallée, Esteban Espinosa, Christophe Noll and Lucie Bouffard for their excellent technical assistance and Stephen C. Cunnane for the use of PMOD. This work was supported by the Québec Network on Drug Research (RQRM), to A.Caron and D.P.Blondin as well as a grant from the Natural Sciences and Engineering Research Council of Canada (NSERC Canada) to D.P.Blondin (RGPIN-2019-05813). D.P. Blondin holds the GSK Chair in Diabetes of the Université de Sherbrooke and a Fonds de Recherche du Québec-Santé (FRQS) J1 salary award. L. Dumont is the recipient of an FRQS doctoral training award. A. Caron is supported by a Canada Research Chair in Neurometabolic Pharmacology and an FRQS J1 salary award. A.C. Carpentier is supported by a Canada Research Chair in the Molecular Imaging of Diabetes.

## Author Contributions

Conceptualization, D.P.B and A.C.; Methodology, L.D., D.P.B, A.C., E.E.T., and A.C.C.; Investigation, L.D., A.C., G.R., E.C., M.F., F.F., S.P., S.D., B.G., E.E.T., A.C.C., and D.P.B.; Writing first draft, L.D. and D.P.B; Writing – review and editing, L.D., A.C., G.R., E.C., M.F., F.F., S.P., S.D., B.G., E.E.T., A.C.C., and D.P.B.; Visualization, L.D. and D.P.B; Funding Acquisition, A.C. and D.P.B.

## Declaration of interests

No conflicts of interest, financial or otherwise, related to this work are declared by the authors.

## STAR METHODS

### KEY RESOURCES TABLE

2 KEY RESOURCES TABLE
| REAGENT or RESOURCE | SOURCE | IDENTIFIER |
| --- | --- | --- |
| **Biological Samples** |  |  |
| Human plasma samples | This paper | N/A |
| Human urine samples | This paper | N/A |
| **Chemicals, Peptides, and Recombinant Proteins** |  |  |
| Myrbetriq (Mirabegron) | Astellas Pharma | N/A |
| Bisoprolol | Sandoz Canada | N/A |
| **Critical commercial Assays** |  |  |
| Glycerol reagent set | Instruchemie, Sigma-Aldrich | Cat# 2913, Cat# F6428 |
| NEFA C kit | Wako diagnostics, Randox laboratories | Cat# 434-91795, Cat# FA115 |
| **Software and Algorithms** |  |  |
| GraphPad Prism 8 for statistical analysis | GraphPad Software | RRID: SCR_002798 |
| Adobe Photoshop | Adobe | https://www.adobe.com/ |
| PMOD 3.7 | PMOD Technologies LLC | RRID : SCR_016547 |
| MATLAB R2019a | The MathWorks | RRID : SCR_001622 |
| **Other** |  |  |
| [<sup>11</sup>C]acetate, radiotracer | CIMS | N/A |
| [<sup>18</sup>F]FDG, radiotracer | CIMS | N/A |
| [6,6-<sup>2</sup>H<sub>2</sub>]-glucose, stable isotope tracer | ACP Chemicals | Cat# 1CDLM349 |
| [U-<sup>13</sup>C]-palmitate, stable isotope tracer | Cambridge isotopes Laboratories | Cat# DLM-1229 |
| [1,1,2,3,3-<sup>2</sup>H<sub>5</sub>]-glycerol, stable isotope tracer | Cambridge isotopes Laboratories | Cat# DLM-1229 |
| Vmax 29n (indirect calorimetry) | Sensormedics | N/A |
| Biograph Vision PET-CT scanner | Siemens Healthineers | N/A |

### RESOURCE AVAILABILITY

#### Lead contact

Further information and requests for resources and reagent should be directed to and will be fulfilled by the lead contact, Denis P. Blondin.

#### Data and code availability

The processed data generated during this study as well as the MATLAB code used to perform the pharmacokinetic modeling of [^18^F]FDG and [^11^C]acetate are available at doi.org/10.5281/zenodo.5834789. Anonymized image files are voluminous and, therefore, not deposited in any public repository, but can be made available on request to the lead contact.

### EXPIRIMENTAL MODEL AND SUBJECT DETAILS

#### Human Subjects

Eight healthy men aged 27 ± 3 years with a body mass index of 22.5 ± 1.7 km/m^2^, body mass of 74.1 ± 3<5.8 kg and body surface area of 2.27 ± 0.09 m^2^ participated in this study. All participants were informed of the methodology and informed written consent was obtained in accordance with the Declaration of Helsinki. The protocol was approved by the Human Ethics Committee of the Centre de recherche du Centre hospitalier universitaire de Sherbrooke and Health Canada provided regulatory approval for the off-label use of mirabegron. The study was pre-registered on ClinicalTrials.gov (NCT04823442). The inclusion criteria included: (i) male; (ii) age between 18 and 35 years; (iii) BMI < 30 kg/m^2^; (iv) normal fasting glucose (< 5.6 mmol/l); (v) normal glucose tolerance (2 h post 75 g OGTT glucose < 7.8 mmol/l); (vi) HbA1c < 5.8%. Exclusion criteria included: (i) overt cardiovascular disease as assessed by medical history, physical exam, and abnormal ECG; (ii) treatment with any drug known to affect lipid or carbohydrate metabolism; (iii) presence of liver disease, uncontrolled thyroid disorder, previous pancreatitis, bleeding disorder, or other major illness; (iv) smoking (>1 cigarette/day) and/or consumption of 2 alcoholic beverages per day; (v) prior history or current fasting plasma cholesterol level > 7 mmol/l or fasting TG > 6 mmol/l.

### METHOD DETAILS

#### Experimental Protocols

Participants took part in two experimental sessions assigned in a random order. Participants ingested either 200 mg of the β_3_-AR agonist mirabegron (Myrbetriq, Astellas Pharma Canada) alone or in combination with 10 mg of bisoprolol, a β_1_-AR antagonist. Each experimental session consisted of a baseline period at ambient temperature (∼22°C), followed by the oral administration of the medication. In the mirabegron + bisoprolol condition, both medications were given simultaneously, as the bisoprolol monogram provide to the FDA reports a t_max_ of 168 min, which is 40 min earlier than the t_max_ of mirabegron (210 min). This would allow us to elicit the effects of β_1_-AR antagonism prior to the full effect of mirabegron-mediated agonism. Experimental sessions were conducted between 08.00 and 16.00 h at the Centre de recherche du Centre hospitalier universitaire de Sherbrooke, following a 12 h fast and 48 h without strenuous physical activity. Participants followed a 2-day standard isocaloric diet which was determined following 3-day food record, accounting for the participant’s standard daily physical activity level, determined by wearing a portable arm band accelerometer for 3-days. Upon arriving to the laboratory, participants emptied their bladder, then were weighed and indwelling catheters were placed in the antecubital vein in both arms for blood sampling and tracer infusion.

#### Metabolic Heat Production, Fuel Selection and Cardiovascular Measures

Whole-body metabolic heat production and substrate utilization was determined by indirect respiratory calorimetry (corrected for protein oxidation), measured every 60 min for a collection period of 20 min ^7^. The molar rate of fatty acid oxidation (µmol·min^-1^) was calculated from triglyceride oxidation (g·min^-1^) assuming the average molecular weight of triglyceride as 861 g·mol^-1^ and multiplying the molar rate of triglyceride oxidation by three, as three moles of fatty acids are contained in each mole of triglyceride. Energy potentials of 16.3, 40.8 and 19.7 kJ·g^-1^ were used to calculate the amount of heat produced from glucose, lipid and protein oxidation, respectively. Blood pressure and heart rate were measured every 60 min using an Omron blood pressure monitor (HEM-790IT, Omron Healthcare).

#### PET Imaging and Analysis

Participants remained supine in a PET and CT scanner (Siemens Healthineers Biograph Vision) for 30 min for the baseline and the final 90 min of the mirabegron treatment. Tissue-specific oxidative metabolism was determined by first performing a CT scan (30 mAs) centered at the cervico-thoracic region to correct for attenuation and define PET regions of interest (ROI). At time -45 min (room temperature, before treatment) and at time 210 min (*i.e.* 210 min following oral administration of mirabegron alone or in combination with bisoprolol), a ∼ 175 MBq bolus of [^11^C]acetate was injected intravenously and was followed by a 20-min list-mode dynamic PET acquisition (frames: 24 x 10 s, 12 x 30 s, 2 x 300 s) centered at the cervico-thoracic junction ^7^. A ∼125 MBq bolus of 2-deoxy-2-[^18^F]fluoro-D-glucose ([^18^F]FDG) was intravenously injected 30 min after the [^11^C]acetate injection at time 240 (*i.e.* 240 min following oral administration of mirabegron alone or in combination with bisoprolol). followed by a list-mode dynamic PET acquisition (frames: 12 x 10 s, 10 x 45 s, 7 x 90 s, 2 x 300 s) centered at the cervico-thoracic junction. Residual [^11^C]acetate activity prior to the [^18^F]FDG scans were corrected by acquiring a 30 s frame prior to the injection of [^18^F]FDG and accounting for the disintegration rate and metabolic clearance of ^11^C.

ROI were drawn from the transaxial CT slices then copied to the [^11^C]acetate and [^18^F]FDG PET images. ROIs were drawn in the left ventricle for blood activity (image-derived arterial input function, AIF), the larger skeletal muscles in the field of view (e.g. *m*. sternocleidomastoid, *m*. trapezius, *m*. pectoralis major, *m*. deltoideus, *m*. levator scapulae, *m*. latissimus dorsi, *m*. erector spinea), on posterior cervical subcutaneous adipose tissue and on supraclavicular BAT. The mean standard uptake values (SUV_mean_) from these ROIs were then extracted for each time frame using PMOD (version 3.7, PMOD Technologies LLC) to create time-activity curves. The blood signal for [^11^C]acetate was then corrected to exclude the contribution of metabolites ^43^. The metabolite- corrected time-activity curves were then used to perform pharmacokinetic modeling in MATLAB (The mathWorks, R2019a). A four-compartment, two-tissue, model^28^ was applied to the [^11^C]acetate signal to derive the rates of uptake (*K*_1_ in ml‧g^-1^‧min^-1^), oxidation (*k*_2_ in min^-1^) and lipid synthesis (*k*_3_ in min^-1^). Plasma and tissue time-radioactivity curves for [^18^F]FDG were analyzed graphically using the Patlak linearization method ^7^. The glucose fractional uptake (*Ki* in min^-1^) is equal to the slope of the plot in the graphical analysis. Net glucose uptake (K_m_ in nmol‧g^-1^‧min^-1^ = (*K*_i_‧circulating glucose)‧(LC^-1^)‧(tissue density‧1000)^-1^) was obtained by multiplying *Ki* by circulating plasma glucose levels at the time of the PET image acquisition, which assumes a lump constant (LC) value of 1.14 and 1.16 compared with endogenous plasma glucose for the BAT and skeletal muscles, respectively, and corrected for a tissue density of 0.925 g/mL and 1.0597 g/mL.

#### Whole-Body Lipolysis and Triacylglycerol/Fatty Acid Cycling

Upon arriving to the laboratory, participants emptied their bladder and indwelling catheters were placed in the antecubital vein in both arms for blood sampling and tracer infusions. A primed (30 µmol/kg) continuous infusion (0.3 µmol·kg^-1^·min^-1^) of [6,6-^2^H_2_]-glucose was started 150 min before oral administration of mirabegron or the combination of mirabegron and bisoprolol (time - 150 min) to determine the systemic rate of appearance of plasma glucose (Ra_glucose_) ^7^. Plasma NEFA appearance rate (Ra_NEFA_) and plasma glycerol appearance rate (Ra_glycerol_) were determined with a continuous infusion of [U-^13^C]-palmitate (0.01 µmol/kg/min in 100 mL 25% human serum albumin) and a primed (1.6 µmol·kg^-1^) continuous infusion (0.05 µmol/kg/min) of [1,1,2,3,3-^2^H_5_]- glycerol started 60 min before the oral administration of mirabegron or the combination of mirabegron and bisoprolol (time - 60 min) ^7^. Ra_NEFA_ was calculated by multiplying the plasma palmitate appearance rate by the fractional contribution of palmitate to total plasma NEFA concentrations.

Total, intracellular and extracellular triacylglycerol/fatty acid (TAG/FA) cycling was calculated using Ra_glycerol_, Ra_NEFA_ and the rate of fatty acid oxidation. In brief, total TAG/FA cycling was calculated as the difference between the rate of fatty acid oxidation and the total amount of fatty acids made available by the hydrolysis of intracellular triglycerides (3 x Ra_glycerol_), which is derived primarily from WAT under fasted conditions. Since glycerol kinase has a low activity in WAT, relative to other tissues, the glycerol produced following the complete hydrolysis of a triglyceride in WAT is rapidly excreted into circulation where it can serve as gluconeogenic substrate. Intracellular or primary TAG/FA cycling refers to the re-esterification of fatty acids within the cell where it was hydrolyzed. This is calculated from the difference between the systemic rate of appearance of NEFA (i.e. Ra_NEFA_) and the total amount of fatty acids released following intracellular hydrolysis of triglycerides (3 x Ra_glycerol_). Extracellular or secondary TAG/FA cycling refers to the re-esterification of fatty acids after it has been released into the circulation, primarily packaged by the liver into triglyceride-rich lipoproteins (very-low density lipoproteins; VLDL) or intracellular lipid droplets. This is calculated from the difference between the total rate of fatty acids oxidation and the systemic appearance of fatty acids (i.e. Ra_NEFA_). The energy cost of TAG/FA cycling was calculated assuming that the re-esterification of each triglyceride requires ∼8 ATP (or ∼144 kcal·mol^-1^ of triglyceride recycled), for the activation of each of the three fatty acids (2 ATP per fatty acid), and glycerogenesis (∼2 ATP) providing the three-carbon backbone for triglyceride synthesis^44^.

#### Biological Assays

Glucose, total NEFA, TG, cortisol, TSH, free T3 and free T4 were measured using specific radioimmunoassay and colorimetric assays ^7^. Plasma C-peptide, GIP, total GLP-1, glucagon, insulin, leptin and PYY were measured using Luminex xMAP-based immunoassays (Milllipore, Etobicoke, ON, Canada). Adiponectin, total and acylated ghrelin were measured by ELISA (Alpco, Salem, NH, USA). Individual plasma NEFA (palmitate, linoleate, oleate), [U-^13^C]- palmitate enrichment, [1,1,2,3,3-^2^H_5_]-glycerol enrichment and [6,6-^2^H_2_]-glucose enrichment were measured by gas chromatography-mass spectrometry ^7^.

### QUANTIFICATION AND STATISTICAL ANALYSIS

#### Statistical Analyses

Statistical analysis was performed using Prism (GraphPad; 8.0). Blood chemistry data are expressed as mean with 95% CI or median with interquartile range otherwise the data are expressed as mean ± SD. The Shapiro-Wilk test was performed to verify the normality of data, when necessary. Significance between groups was determined using 2-tailed paired Student’s t test, or one-way or two-way ANOVA when appropriate, with multiple comparisons using Bonferroni’s correction. The significance threshold was set at P ≤ 0.05.

### ADDITIONAL RESOURCES

ClinicalTrials.gov identifier NCT04823442

## Notes

Conflict of interest: The authors have declared that no conflict of interest exists related to the content of this manuscript

### Competing Interest Statement

The authors have declared no competing interest.

### Clinical Trial

NCT04823442

### Funding Statement

This work was supported by the Quebec Network on Drug Research (RQRM), to A.Caron and D.P.Blondin as well as a grant from the Natural Sciences and Engineering Research Council of Canada (NSERC Canada) to D.P.Blondin (RGPIN-2019-05813). D.P. Blondin holds the GSK Chair in Diabetes of the Universite de Sherbrooke and a Fonds de Recherche du Quebec-Sante (FRQS) J1 salary award. L. Dumont is the recipient of an FRQS doctoral training award. A. Caron is supported by a Canada Research Chair in Neurometabolic Pharmacology and an FRQS J1 salary award. A.C. Carpentier is supported by a Canada Research Chair in the Molecular Imaging of Diabetes

### Author Declarations

Ethics committee of the CIUSSS de l'Estrie - CHUS gave ethical approval for this work.

